# Four longitudinal phenotypes of radiation-associated dysphagia following oropharyngeal radiotherapy: a latent class trajectory analysis

**DOI:** 10.64898/2026.07.06.26357052

**Authors:** Beatrice Manduchi, Carly E. A. Barbon, Amy C. Moreno, Christine B. Peterson, David Swanson, Jack J Lee, Anna Lee, Andrew J. Schaefer, Clifton D. Fuller, Stephen Y. Lai, Steven J. Frank, Katherine A. Hutcheson, the OPC-SURVIVOR Research program

## Abstract

**Background and purpose:** Patients with oropharyngeal cancer (OPC) treated with radiotherapy (RT) exhibit heterogeneous courses of radiation-associated dysphagia (RAD) during recovery, yet most survivorship models typically treat RAD uniformly. This study aimed to identify distinct, data-driven RAD longitudinal Phenotypes based on imaging-graded swallow function from pre-treatment to 30 months post-RT and to characterize their baseline predictors.

**Materials and methods:** Heterogeneous linear mixed-effects latent class trajectory modeling was applied to longitudinal DIGEST scores from the Stiefel MDA-OPC prospective registry. Eligible patients had ≥3 Modified Barium Swallow (MBS) assessments between baseline and 30 months post-RT. Models were evaluated across functional forms and 1-5 latent classes; final selection used the Bayesian Information Criterion. Baseline predictors of class membership were identified via binary logistic regression.

**Results:** The cohort comprised 650 OPC patients (2,116 MBS assessments; mean age 61 years, 89% male, 93% HPV-positive). Four RAD Phenotypes were identified: No/Minimal RAD (n=385/650, 59%), Mild/Moderate RAD (n=104/650, 16%), Moderate/Severe Transient RAD (n=94/650, 15%), and Moderate/Severe Progressing RAD (n=67/650, 10%). Classification quality was acceptable (mean posterior probabilities 0.78-0.89; entropy 0.69). Baseline DIGEST impairment, base-of-tongue primary, advanced T stage, and age ≥60 independently predicted membership in higher-burden Phenotypes (AUC=0.845; 10-fold CV-AUC=0.835).

**Conclusion:** RAD following RT for OPC comprises four biologically and clinically distinct longitudinal Phenotypes, predictable from pre-treatment characteristics. These findings support trajectory phenotyping as outcome framework for RAD research and risk-adaptive survivorship care.

## INTRODUCTION

Oropharyngeal cancer (OPC) has undergone a marked epidemiologic shift, driven by the rising incidence of human papillomavirus (HPV)-associated disease^1^. OPC is a common cancer in the United States, with roughly 54,000 new cases diagnosed annually and a predominance of HPV-positive tumors that affect younger, otherwise healthier adults(Siegel et al., 2025). This epidemiologic transition has yielded survival probabilities that exceed 90% at two years and approach 75% at five years^2–4^, redefining clinical priorities toward long-term functional outcomes and quality of life. Despite advances in radiotherapy (RT) and supportive care that have mitigated acute morbidity, chronic treatment-related toxicities remain pervasive and clinically consequential across survivorship.

Radiation-associated dysphagia (RAD) stands as one of the most common and functionally disabling late effect of RT for OPC. Population models demonstrate that dysphagia severity peaks early post-treatment and improves on average within the first six months^5,6^, yet a meaningful proportion of patients experience persistent or delayed clinically significant swallowing dysfunction and its clinical consequences beyond two years^7–10^. This pattern, observed across diverse outcomes including patient-reported and clinician-graded metrics (e.g., aspiration prevalence, image-based DIGEST or CTCAE), underscores the enduring burden of RAD even in modern cohorts.

Despite this recognition, the prevailing analytic framework for RAD reduces this experience to population-mean trajectories (i.e., an acute peak followed by partial recovery) implying homogeneity in development and recovery patterns^5^. This assumption is inconsistent with clinical reality: some survivors preserve swallowing throughout treatment and recovery, while others plateau with persistent low/mid-grade impairment, and others deteriorate progressively through mechanisms including fibrosis and neurovascular compromise from cumulative radiobiological injury^11–13^. The determinants of these divergent courses, including radiation dose to swallowing-related structures, tumor subsite, concurrent chemotherapy, and host factors, are well established in NTCP models^14,15^, yet their integration into individualized trajectory prediction remains limited.

The clinical stakes of this gap are consequential. A one-size-fits-all approach to dysphagia surveillance and rehabilitation may misalign resources, over-surveilling some and missing opportunities for earlier, targeted intervention in others. Patients with durably preserved swallowing may be subjected to overly intensive monitoring, while those at risk for progressive impairment may receive insufficient interventions to preserve function and mitigate complications such as aspiration, malnutrition, and social withdrawal. Consequently, current risk models do not predict individual recovery trajectories, limiting their utility in shared decision-making and personalized care planning.

Recognition of heterogeneous disease and symptom trajectories has led to increasing adoption of longitudinal phenotyping approaches across clinical and oncologic research. Rather than summarizing outcomes using a single population-average trajectory, latent class and growth mixture modeling techniques identify subgroups of individuals who share qualitatively similar patterns of change over time, capturing clinically meaningful heterogeneity that conventional analyses may obscure^16^. These approaches have been successfully applied to characterize diverse trajectories of symptoms, functional outcomes, and recovery, demonstrating that subgroup-specific patterns often possess distinct prognostic and therapeutic significance beyond cross-sectional or mean-based estimates^17,18^.

In RAD, Christianen et al. provided early evidence of this heterogeneity by applying cluster analysis to clinician-rated swallowing dysfunction assessed at four timepoints up to 24 months post-RT in a prospective cohort of 238 head and neck cancer patients^19^. Five distinct patterns emerged, each associated with different dose distributions to swallowing organs at risk, leading the authors to hypothesize that these patterns reflect distinct underlying radiobiological processes, including reversible edema, progressive fibrosis, and recovery of salivary flow. However, that work was conducted in a mixed head and neck cancer population using a clinician-rated toxicity scale, which lack the sensitivity and mechanistic granularity of instrumental swallowing assessment. Whether analogous phenotypes exist specifically in OPC, a disease site with distinct anatomy, treatment volumes, and a now-dominant HPV-associated biology, and whether they can be identified and characterized using objective, image-based measures of swallowing physiology, remains unknown. Addressing these gaps, this study uses longitudinal, image-graded swallowing assessments to: 1) identify and classify distinct, longitudinal phenotypes of RAD in the first 30 months after RT; 2) evaluate their associations with baseline clinical and treatment-related factors; and 3) validate these Phenotypes in an independent multisite cohort. Such Phenotypes could provide a basis for personalized dysphagia surveillance, risk-adaptive prehabilitation, and more precise survivorship care frameworks tailored to the heterogeneous reality of functional recovery after RT.

## METHODS

### Study design and participants

This was a retrospective analysis of prospectively collected institutional registry, the Stiefel MDA-OPC cohort^20^ (Institutional protocol: PA14-0947), a longitudinal cohort of OPC patients with standardized clinical, treatment, and patient-reported measures at defined intervals from pre-treatment through survivorship. Eligible patients had a confirmed diagnosis of OPC, received primary or adjuvant RT (± concurrent chemotherapy), and had at least three MBS studies within the study window: one baseline assessment (performed within six months prior to RT start; when multiple baselines were available, the one closest to RT start was retained) and at least two post-RT assessments within 30 months of RT completion. The analytic dataset was supplemented with off-protocol clinical MBS studies when available. This study involved human participants and was approved by The University of Texas MD Anderson Cancer Center Institutional Review Board (Protocol PA14-0947), which includes approval for secondary analyses.

### Swallow imaging assessment

MBS studies were performed according to the Radiation Swallowing Care Pathway (“RADPath”) at pre-specified timepoints regardless of dysphagia symptoms (pre-RT, 3–6 months post-RT, and 18–24 months post-RT); additional MBS were ordered as clinically indicated (i.e., symptomatic referrals). All assessments were conducted using a standardized protocol by a trained speech-language pathologist (SLP)^21^, and recorded at 15–30 frames per second using the TIMS DICOM System (Foresight Imaging LLC). The protocol included thin liquid trials (5 cc, 10 cc, cup sip), barium pudding, and crackers (Varibar®, Bracco Diagnostics, Inc.). Each MBS was scored using the Dynamic Imaging Grade of Swallowing Toxicity (DIGEST) scale^22^, a validated, MBS-based instrument aligned with the NCI Common Terminology Criteria for Adverse Events, providing a summary rating of pharyngeal swallowing function integrating safety and efficiency profiles (grade 0 = no dysphagia, 1 = mild, 2 = moderate, 3 = severe, 4 = life-threatening). Safety is determined by the pattern and severity of airway invasion per the Penetration-Aspiration Scale (PAS)^23^; efficiency reflects ordinal estimates of pharyngeal residue. All studies were laboratory-graded by SLPs trained to reliability standards (≥80% exact agreement with a gold-standard rater).

### Clinical and patient-reported measures

Demographic, disease, and treatment characteristics were extracted from the prospective registry. Baseline patient-reported swallowing-related quality of life was assessed using the MD Anderson Dysphagia Inventory (MDADI), a validated 20-item instrument measuring the physical, functional, and emotional impact of dysphagia on quality of life (composite score 0–100; higher scores indicate better function)^24^. Oral intake at baseline was assessed using the Performance Status Scale for Head and Neck Cancer (PSS-HN) Normalcy of Diet subscale, a clinician-rated measure of normalcy of diet (score 0–100; higher scores indicate less dietary restriction)^25^.

### Statistical analysis

Distribution of sample characteristics and outcome of interests were reported descriptively, using means (with SD) and frequencies (with %).

#### Aim 1: Phenotypes identification

To identify longitudinal Phenotypes of RAD, heterogeneous linear mixed-effects (HLME) latent class trajectory modelling was applied using the lcmm package in R^26^. DIGEST (outcome) and months since RT completion (predictor) were treated as a continuous. Four functional forms were evaluated: linear, quadratic polynomial, and natural cubic splines with 3 or 4 degrees of freedom (df; with automatic knot placement). For each functional form, models with 1 to 5 latent classes were fitted. Class-specific residual variance was permitted via proportional scaling, and a random intercept was included in all models. Models were considered inadmissible if any class contained fewer than 10% of the analytic sample^16^. Among admissible solutions, the model with the lowest Bayesian Information Criterion (BIC) was selected^27^. Classification quality was assessed via mean posterior probabilities of assigned class membership (threshold ≥0.70) and relative entropy (range 0–1; higher values indicate cleaner separation)^16^.

#### Aim 2: Phenotype characterization and baseline prediction model

Baseline characteristics were compared across longitudinal Phenotypes using Chi-square or Fisher’s exact tests for categorical variables and independent-samples t-tests or Mann-Whitney U tests for continuous variables. Pairwise comparisons with Bonferroni correction were performed across all Phenotypes pairs. To predict Phenotype membership at baseline, an exploratory binary logistic regression model was developed contrasting the lowest-risk Phenotype (Class 0) against all higher-risk Phenotypes combined (Classes 1–3). Candidate predictors were variables reaching statistical significance in univariate pairwise comparisons and were dichotomized using clinically grounded thresholds (e.g., age at 60 years, the cohort median; MDADI at 74, threshold for impaired swallowing-related QOL^28^; baseline DIGEST 0 vs ≥1). Model discrimination was assessed by the area under the receiver operating characteristic curve (AUC, 95% CI), with internal validity assessed via 10-fold cross-validated AUC. Results are expressed as odds ratios (ORs) with 95% CIs. Two pre-specified sensitivity analyses were conducted: (1) replacing dichotomization of baseline DIGEST as 0-1 vs ≥2; and (2) replacing dichotomized predictors with continuous variables. All analyses were performed in R (version 2025.09.1+401). Statistical significance was set at two-sided p<0.05.

## RESULTS

The cohort included 650 patients with 2,116 MBS studies (Table 1). Mean age was 61.0 years (SD 8.5); the majority were male (578/650, 88.9%) and HPV/p16-positive (595/650, 92.5%), with a roughly equal split between base of tongue and tonsil primaries (49.5% and 42.3%, respectively). Most patients (498/650, 76.6%) had three MBS assessments, 138/650 (21.2%) had four, and 14/650 (2.2%) had five. Of 2,116 MBS studies, 650 (30.7%) were conducted at pre-treatment, 583 (27.6%) at 1–6 months, 186 (8.8%) at 6–12 months, 205 (9.7%) at 12–18 months, and 492 (23.3%) at 18–30 months post-RT (Supplementary eFigure 1 shows DIGEST distribution per time period).

**Table 1.** Pre-treatment cohort characteristics, overall and by longitudinal RAD Phenotype (n=650)

|  | Total cohort | Longitudinal RAD Phenotypes |  |  |  | p-value* |
| --- | --- | --- | --- | --- | --- | --- |
|  |  | (0) No/<br>Minimal<br>RAD | (1) Mild/<br>Moderate<br>RAD | (2) Moderate/<br>Severe<br>Transient<br>RAD | (3) Moderate/<br>Severe<br>Progressing<br>RAD |  |
| **N (%)** | 650 (100%) | 385 (59.2%) | 104 (16.0%) | 94 (14.5%) | 67 (10.3%) |  |
| **Age, mean (SD)** | 61.0 (8.5) | 59.2 (8.1) | 63.9 (7.7) | 63.8 (8.6) | 63.3 (9.2) | **<0.001** |
| **Sex** |  |  |  |  |  |  |
| Female | 72 (11.1%) | 48 (12.5%) | 11 (10.6%) | 8 (8.5%) | 5 (7.5%) | 0.513 |
| Male | 578 (88.9%) | 337 (87.5%) | 93 (89.4%) | 86 (91.5%) | 62 (92.5%) |  |
| **Tumor site** |  |  |  |  |  |  |
| BOT | 322 (49.5%) | 145 (37.7%) | 74 (71.2%) | 56 (59.6%) | 47 (70.1%) | **<0.001** |
| Tonsil | 275 (42.3%) | 201 (52.2%) | 25 (24.0%) | 32 (34.0%) | 17 (25.4%) |  |
| Other | 53 (8.2%) | 39 (10.1%) | 5 (4.8%) | 6 (6.4%) | 3 (4.5%) |  |
| **HPV/ p16 status** |  |  |  |  |  |  |
| Negative | 48 (7.5%) | 23 (6.0%) | 8 (7.8%) | 9 (9.7%) | 8 (12.3%) | 0.258 |
| Positive | 595 (92.5%) | 359 (94.0%) | 95 (92.2%) | 84 (90.3%) | 57 (87.7%) |  |
| **T stage** |  |  |  |  |  |  |
| T0 | 35 (5.4%) | 27 (7.0%) | 4 (3.8%) | 2 (2.1%) | 2 (3.0%) | **<0.001** |
| T1 | 190 (29.2%) | 140 (36.4%) | 19 (18.3%) | 16 (17.0%) | 15 (22.4%) |  |
| T2 | 235 (36.2%) | 144 (37.4%) | 37 (35.6%) | 40 (42.6%) | 14 (20.9%) |  |
| T3 | 111 (17.1%) | 59 (15.3%) | 16 (15.4%) | 20 (21.3%) | 16 (23.9%) |  |
| T4 | 79 (12.2%) | 15 (3.9%) | 28 (26.9%) | 16 (17.0%) | 20 (29.9%) |  |
| **N stage** |  |  |  |  |  |  |
| N0 | 52 (8.0%) | 25 (6.5%) | 7 (6.7%) | 15 (16.0%) | 5 (7.5%) | **<0.001** |
| N1 | 423 (65.1%) | 285 (74.0%) | 50 (48.1%) | 50 (53.2%) | 38 (56.7%) |  |
| N2 | 159 (24.5%) | 65 (16.9%) | 44 (42.3%) | 27 (28.7%) | 23 (34.3%) |  |
| N3 | 16 (2.5%) | 10 (2.6%) | 3 (2.9%) | 2 (2.1%) | 1 (1.5%) |  |
| **RT type** |  |  |  |  |  |  |
| VMAT | 356 (54.8%) | 210 (54.5%) | 61 (58.7%) | 56 (59.6%) | 29 (43.3%) | 0.429 |
| IMRT | 144 (22.2%) | 85 (22.1%) | 21 (20.2%) | 17 (18.1%) | 21 (31.3%) |  |
| IMPT | 148 (22.8%) | 89 (23.1%) | 21 (20.2%) | 21 (22.3%) | 17 (25.4%) |  |
| Other | 2 (0.4%) | 1 (0.3%) | 1 (1.0%) | 0 (0.0%) | 0 (0.0%) |  |
| **RT dose (Gy), mean** | 68.2 (3.8) | 68.2 (3.9) | 67.6 (4.7) | 68.6 (2.9) | 68.8 [2.60 |  |
| **(SD) [min; max]** | [18.9; 71.1] | [18.9; 70.7] | [42.4; 71.1] | [55.1; 70.4] | [60.0; 70.8] | 0.313 |
| **RT dose (binary)** |  |  |  |  |  |  |
| <66 Gy | 77 (11.8%) | 45 (11.7%) | 16 (15.4%) | 9 (9.6%) | 7 (10.4%) | 0.605 |
| ≥66 Gy | 573 (88.2%) | 340 (88.3%) | 88 (84.6%) | 85 (90.4%) | 60 (89.6%) |  |
| **Concurrent chemotherapy** |  |  |  |  |  |  |
| No | 166 (25.5%) | 111 (28.8%) | 25 (24.0%) | 17 (18.1%) | 13 (19.4%) | 0.094 |
| Yes | 484 (74.5%) | 274 (71.2%) | 79 (76.0%) | 77 (81.9%) | 54 (80.6%) |  |
| **Surgery (primary)** |  |  |  |  |  |  |
| No | 593 (91.2%) | 349 (90.6%) | 90 (86.5%) | 90 (95.7%) | 64 (95.5%) | 0.073 |
| Yes | 57 (8.8%) | 36 (9.4%) | 14 (13.5%) | 4 (4.3%) | 3 (4.5%) |  |
| **Baseline MDADI** |  |  |  |  |  |  |
| **Composite, mean (SD)** | 87.8 (13.4) | 90.3 (11.5) | 80.7 (17.1) | 87.5 (12.7) | 84.0 (14.8) |  |
| **[min; max]** | [24.2; 100] | [24.2; 100] | [28.4; 100] | [47.4; 100] | [44.2; 100] | **<0.001** |
| **Baseline PSS-HN Diet** |  |  |  |  |  |  |
| Non-oral (0) | 2 (0.3%) | 0 (0.0%) | 2 (2.2%) | 0 (0.0%) | 0 (0.0%) | **<0.001** |
| Liquid (10-20) | 3 (0.5%) | 0 (0.0%) | 1 (1.1%) | 2 (2.4%) | 0 (0.0%) |  |
| Pureed/ non/chewable (30-40) | 8 (1.3%) | 2 (0.6%) | 3 (3.2%) | 0 (0.0%) | 3 (5.0%) |  |
| Restricted solids (50-80) | 65 (10.9%) | 28 (7.8%) | 19 (20.4%) | 11 (13.3%) | 7 (11.7%) |  |
| Full, liquid assist (90) | 41 (6.9%) | 16 (4.5%) | 13 (14.0%) | 7 (8.4%) | 5 (8.3%) |  |
| Full, no restriction (100) | 476 (80.0%) | 313 (87.2%) | 55 (59.1%) | 63 (75.9%) | 45 (75.0%) |  |
| **Baseline DIGEST** |  |  |  |  |  |  |
| 0 | 460 (70.8%) | 344 (89.4%) | 9 (8.7%) | 62 (66.0%) | 45 (67.2%) | **<0.001** |
| 1 | 157 (24.2%) | 38 (9.9%) | 66 (63.5%) | 32 (34.0%) | 21 (31.3%) |  |
| 2 | 24 (3.7%) | 3 (0.8%) | 20 (19.2%) | 0 (0.0%) | 1 (1.5%) |  |
| 3 | 8 (1.2%) | 0 (0.0%) | 8 (7.7%) | 0 (0.0%) | 0 (0.0%) |  |
| 4 | 1 (0.2%) | 0 (0.0%) | 1 (1.0%) | 0 (0.0%) | 0 (0.0%) |  |

\* Kruskal-Wallis test for continuous variables; Pearson chi-square test for categorical variables
Legend: BOT, base of tongue; DIGEST, Dynamic Imaging Grade of Swallowing Toxicity; IMPT, intensity-modulated proton therapy; IMRT, intensity-modulated radiation therapy; MDADI, MD Anderson Dysphagia Inventory; PSS-HN, Performance Status Scale for Head and Neck Cancer; RAD, radiation-associated dysphagia; RT, radiation therapy; VMAT, volumetric modulated arc therapy.

### Longitudinal RAD Phenotypes (Aim 1)

Across 16 candidate models (Supplementary eTable 1), the natural cubic spline (df=3) with four latent classes was selected on the basis of the lowest BIC among all admissible solutions (BIC=4548.1; full parameter estimates in Supplementary eTable 2). The final model identified four distinct RAD Phenotypes (Figure 1): **(0) No/Minimal RAD** (n=385/650, 59.2%), characterized by persistently low DIGEST scores approaching 0 with minimal change across all times (pre and post-RT); **(1) Mild/Moderate RAD** (n=104/650, 16.0%), with baseline dysphagia approaching grade 1 (mild) and stable mild/moderate dysfunction post-RT; **(2) Moderate/Severe Transient RAD** (n=94/650, 14.5%), characterized by pronounced acute worsening early post-RT, followed by partial subacute recovery; **(3) Moderate/Severe Progressing RAD** (n=67/650, 10.3%), with progressive worsening across acute and subacute phases, stabilizing at a high severity level in the late follow-up period. Supplementary eTable 3 shows cumulative change in predicted DIGEST grade by Phenotype and time-period.

**Figure 1.**
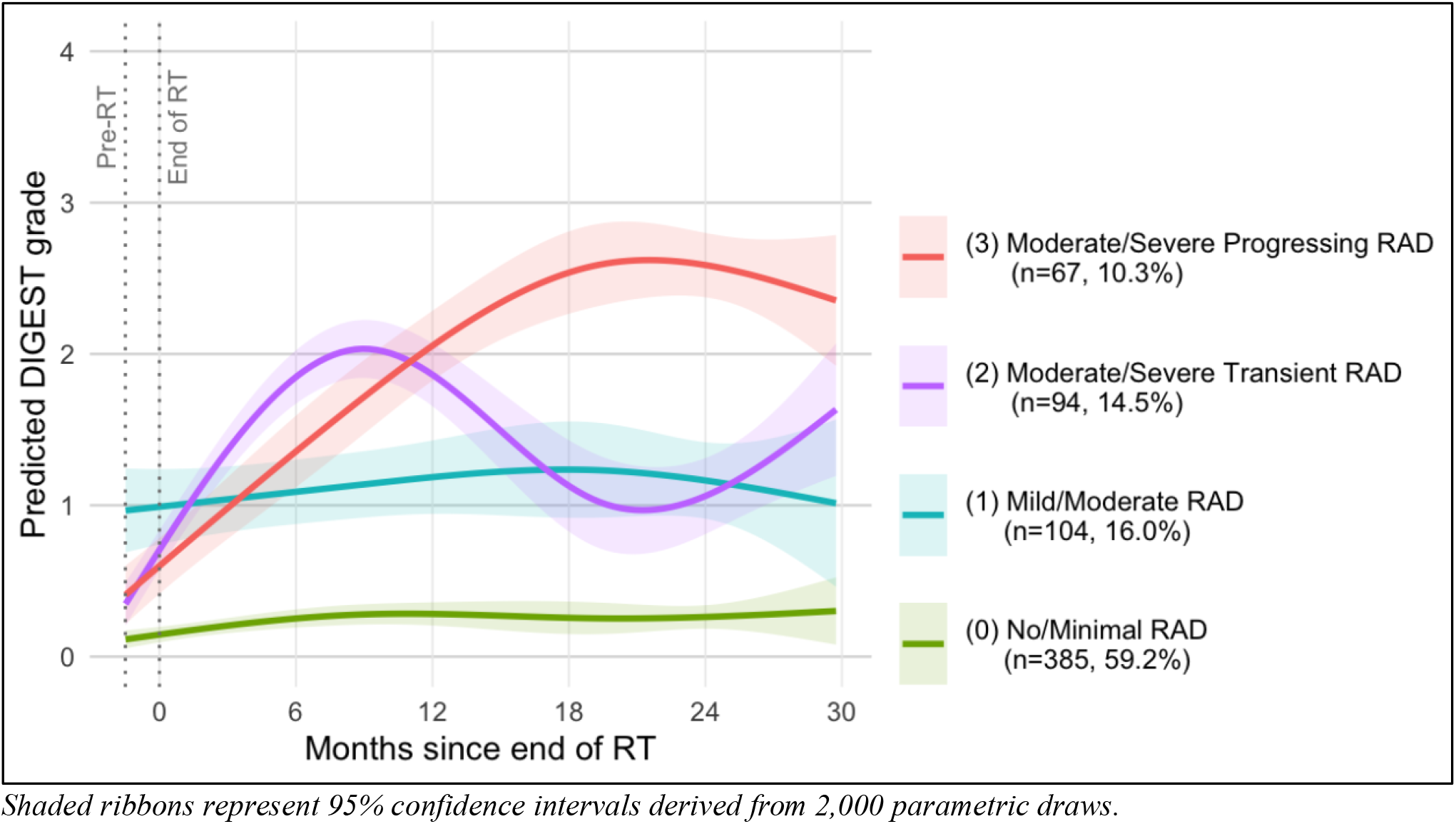
The four longitudinal RAD Phenotypes: Model-predicted DIGEST grade trajectories with 95% confidence intervals for four latent classes identified by heterogeneous linear mixed-effects modelling with natural cubic spline trajectory (df=3) over 30 months following radiotherapy (n=650 patients; 2,116 MBS assessments).

Classification quality was acceptable across all Phenotypes: mean posterior probabilities of assigned Phenotype membership ranged from 0.78 to 0.89 (all exceeding the pre-specified threshold of 0.70), and overall relative entropy was 0.69 (Supplementary eTable 2). Model stability was confirmed by refitting with an independent seed (seed=999), which reproduced the primary solution exactly (BIC=4548.1; identical Phenotype distribution). Individual observed DIGEST trajectories by Phenotype are shown in Supplementary eFigure 2.

### Baseline Phenotypes characteristics and exploratory prediction model (Aim 2)

Baseline characteristics differed significantly across the four RAD Phenotypes for age, tumor site, T/N stage, baseline MDADI composite, PSS-HN diet, and DIGEST grade (Table 1, Supplementary eTable 3). Phenotype 0 was youngest (mean 59.2 years), tonsil-predominant (52%), early T stage (T0–T2: 81%), with near-normal baseline swallowing (DIGEST 0: 89%; full diet: 87%; MDADI: 90.3). Phenotype 1 was characterized by markedly impaired baseline swallowing (91% had DIGEST ≥1 at baseline, 28% DIGEST ≥2, mean MDADI was 80.7, and only 59% reported a full unrestricted diet), despite tumor burden broadly similar to the higher-severity Phenotypes. In contrast, Phenotypes 2 and 3 had largely preserved baseline swallowing function (DIGEST 0: 66% and 67%; MDADI: 87.5 and 84.0) in the setting of more advanced nodal disease (N2–N3: 31% and 36%), and predominantly BOT primaries (60% and 70%).

Pairwise comparisons (Figure 2) revealed differences were predominantly driven by Phenotype 0 versus all higher-risk ones. Significant risk differences between Phenotypes 1, 2, and 3, were limited to prior surgery (Phenotype 1 vs. 2, p=0.024) and baseline DIGEST, which differed across all pairs except Phenotypes 2 vs 3.

**Figure 2.**
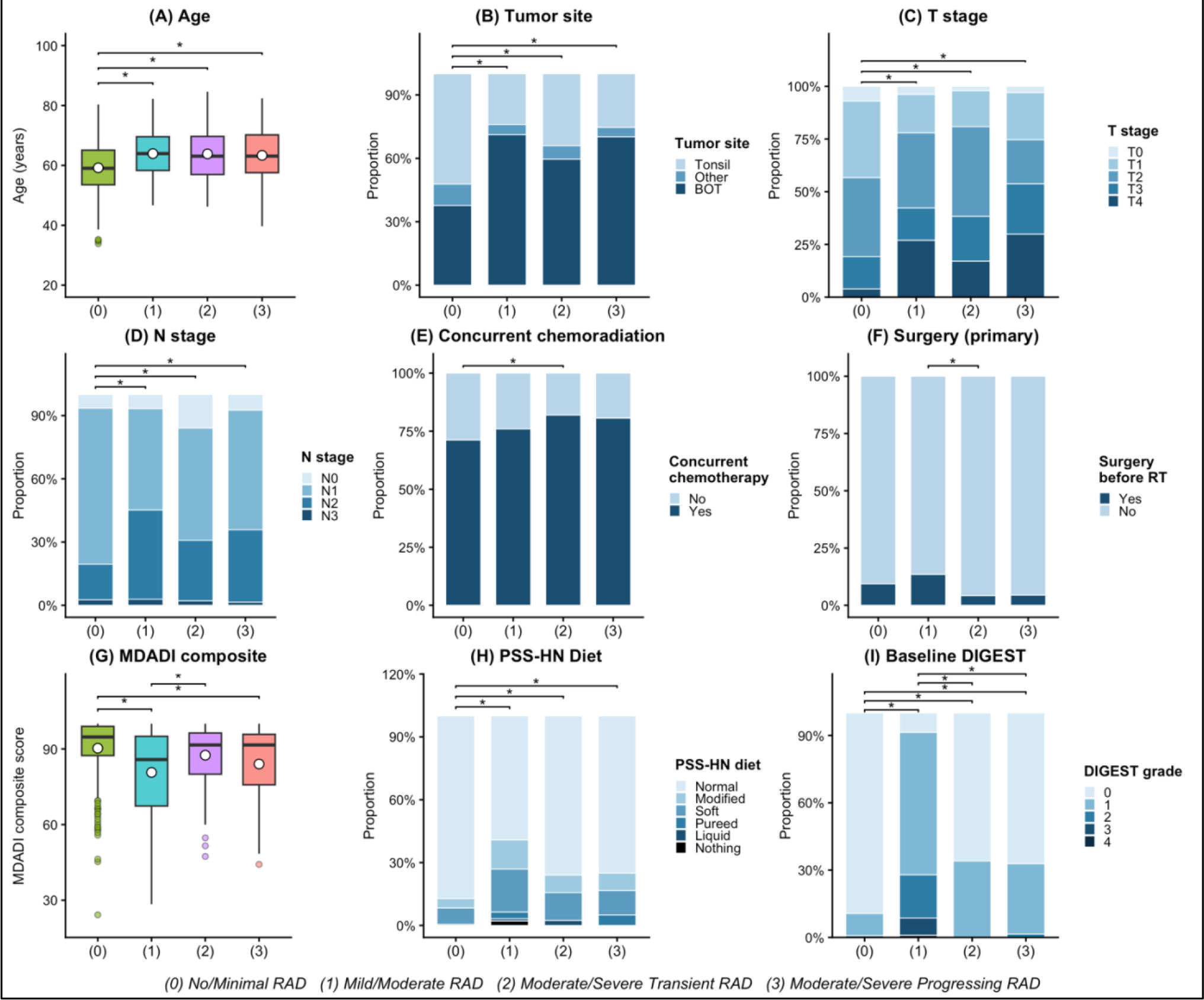
Pre-treatment characteristics across RAD Phenotypes (n=650). Note: *indicates p<0.05 for pairwise comparisons. Kruskal-Wallis with Dunn’s post-hoc correction for continuous variables; Pearson chi-square for categorical variables. Legend: BOT, base of tongue; CT, chemotherapy; DIGEST, Dynamic Imaging Grade of Swallowing Toxicity; MDADI, MD Anderson Dysphagia Inventory; PSS-HN, Performance Status Scale for Head and Neck Cancer; RAD, radiation-associated dysphagia; RT, radiation therapy.

The binary logistic regression model predicted probability of Phenotype 1–3 membership vs Phenotype 0 in n=521 patients with complete predictor data (Figure 3). Independent predictors of higher-risk RAD Phenotypes were: any baseline swallowing impairment (DIGEST ≥1 vs. 0), BOT tumor site (vs. tonsil), advanced T stage (T3–4 vs. T0–2), and age ≥60 years. The model demonstrated good discrimination (AUC=0.85, 95% CI: 0.81–0.88; 10-fold CV-AUC=0.84) and adequate calibration (Hosmer–Lemeshow χ^2^=12.18, df=8, p=0.144). Both sensitivity analyses yielded consistent predictor profiles and comparable discrimination (Sensitivity 1: AUC=0.78, CV-AUC=0.76; Sensitivity 2: AUC=0.85, CV-AUC=0.84).

**Figure 3.**
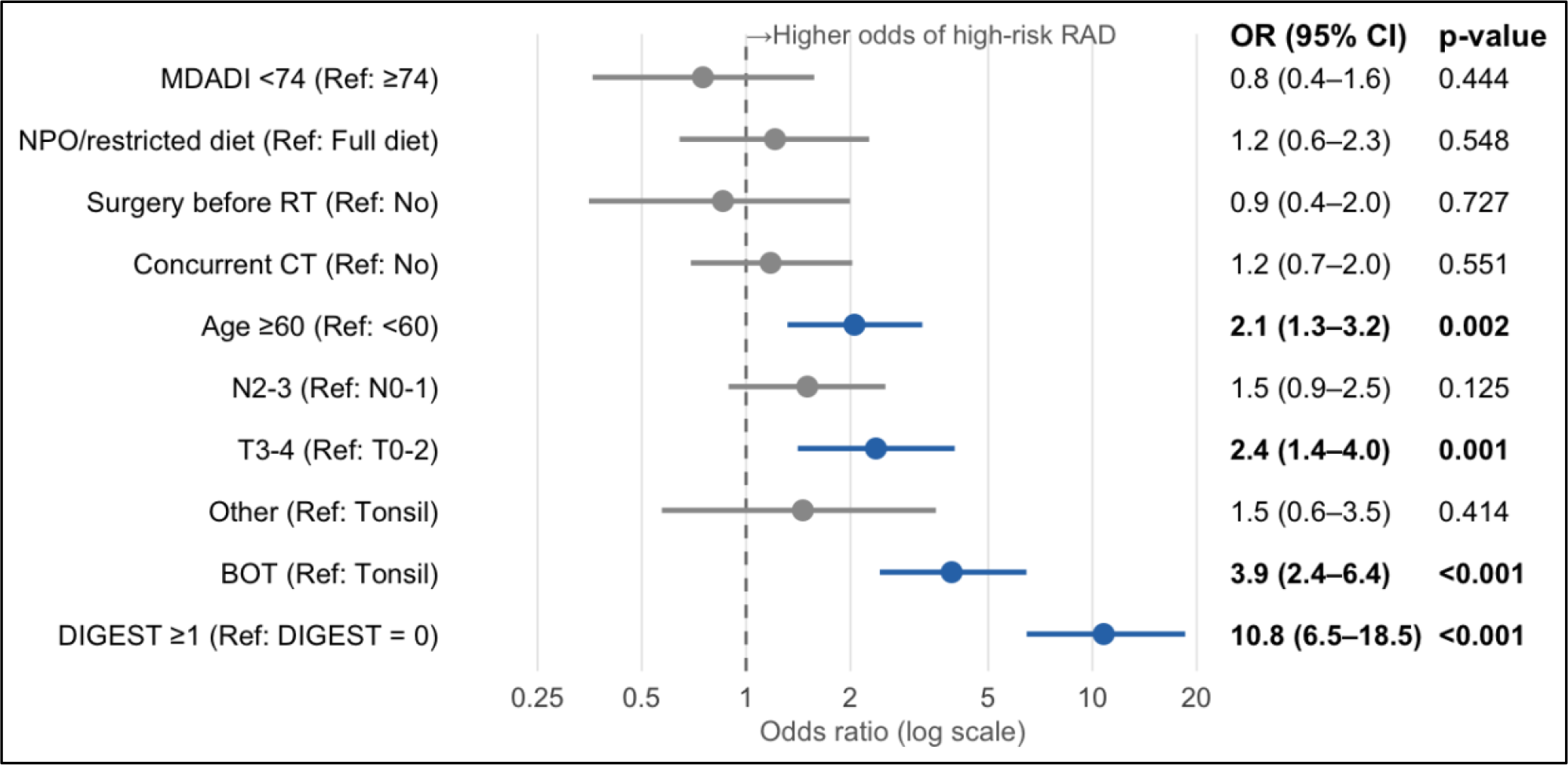
Baseline multivariable logistic regression model predicting membership in a high-risk RAD Phenotypes (1–3) versus low-risk RAD Phenotype (0) at baseline (n=521). Blue points indicate statistically significant predictors (p<0.05). Legend: OR, odds ratio; CI, confidence interval; BOT, base of tongue; CT, chemoradiotherapy; DIGEST, Dynamic Imaging Grade of Swallowing Toxicity; baseline MDADI, MD Anderson Dysphagia Inventory; NPO = nihil per os (nothing by mouth); RAD, radiation-associated dysphagia; RT, radiotherapy.

## DISCUSSION

In the contemporary HPV-associated OPC era, where long-term survival is now the expected outcome for most patients, RAD is regarded as one of the most clinically consequential survivorship challenges of RT. Existing literature has largely characterized RAD through population-mean trajectories or cross-sectional endpoints, approaches that assume homogeneity in progression and recovery, and risk obscuring meaningful interindividual variability. In this study, we applied latent trajectory modeling to longitudinal, image-graded swallowing assessments in a large prospective OPC cohort and identified four distinct longitudinal RAD Phenotypes over 30 months post-RT, demonstrating that swallowing recovery is heterogeneous. Distinct longitudinal Phenotypes observed and validated here thereby demonstrate the clinical course is inadequately captured by group-average estimates alone.

The identified Phenotypes appeared to reflect distinct clinical and biological profiles rather than a simple severity continuum. Almost 60% of patients were in the ‘ideal’ swallow Phenotype (No/Minimal RAD) with near normal baseline swallow (DIGEST grade 0) preserved through 30 months; this Phenotype was younger, more likely to have tonsillar primaries and lower T/N stage disease, consistent with a population with greater functional reserve, low-risk disease, and lower inherent susceptibility to treatment-related injury. The Mild/Moderate Phenotype was primarily distinguished by markedly impaired baseline swallowing function (concentrating nearly all pre-treatment DIGEST ≥2 cases in the cohort) with lower MDADI scores prior to any treatment. These patients were also older and had higher rates of prior surgery, suggesting that pre-existing functional compromise rather than acute treatment injury may be the principal driver of this Phenotype. In contrast, the Moderate/Severe Transient and Progressing ones had generally preserved baseline function despite advanced N stage and concurrent chemoradiotherapy, implicating treatment-mediated injury as the dominant mechanism (through larger radiation fields, higher dose exposure to swallowing-related structures, and intensified systemic therapy). Taken together, these patterns suggest that high-risk RAD may emerge from at least two distinct injury mechanisms: pre-treatment functional impairment (i.e., tumor, comorbidity) shaping the Mild/Moderate Phenotypes, and treatment-mediated acute injury driving the Moderate/Severe ones, which then diverge in their capacity for recovery. This mechanistic interpretation is biologically plausible. The Moderate/Severe Transient Phenotype (characterized by marked acute worsening followed by recovery) is consistent with reversible inflammatory mucosal injury, edema, and transient neuromuscular dysfunction ^13^. Conversely, the Progressing Phenotype, which exhibited continued worsening with minimal late recovery, raises the possibility of progressive fibrosis, muscular atrophy, neuropathy, or vascular compromise of swallowing-related structures^29,30^. Whether this represents true late-radiation syndrome or delayed stabilization will require follow-up beyond 30 months to determine.

These findings should be interpreted in the clinical context in which they were derived. The cohort originated from a high-volume tertiary cancer center employing an intensive, proactive swallowing care model (including standardized serial MBS surveillance and early speech-language pathology involvement throughout treatment and into survivorship). The observation that nearly 60% of patients maintained near-normal swallowing function may thus reflect, in part, what intensive multidisciplinary swallowing care can achieve. Conversely, the fact that 40% of patients in a high intensity swallow preservation environment still develop or persist with meaningful degrees of RAD could also be interpreted as an opportunity for innovative solutions to swallow preservation to extend popular prehabilitation models.

Although prior work has recognized heterogeneity in swallowing outcomes after RT, relatively few studies have explicitly modeled longitudinal recovery trajectories. As previously mentioned, Christianen et al. (2015) identified five distinct patterns of radiation-associated swallowing dysfunction in a heterogeneous head and neck cancer cohort using cluster analysis of clinician-rated toxicity, demonstrating differential swallowing organs-at-risk dosimetry associated with trajectory patterns^19^. Namely, laryngeal dose related to acute dysphagia then attenuated whereas pharyngeal dose durably associated with dysphagia through 2 years. That foundational work was, however, limited by reliance on clinician-rated severe toxicity rather than imaging, a mixed-site population dominated by laryngeal primaries, and exclusion of patients with any abnormal baseline swallow function. The present study extends this literature by applying HLME trajectory modeling to dense MBS-based assessment DIGEST data in a large, OPC-specific survivorship cohort, enabling more flexible characterization of trajectory shape and formal external validation. Notably, despite population and methodological differences, the broad Phenotype typology identified here conceptually parallels the patterns described by Christianen and colleagues (low-persistent, moderate-persistent, transient, progressive and severe-persistent), supporting the biological reproducibility of these recovery archetypes across broader HNC contexts.

The exploratory baseline prediction model provides an initial step toward clinically actionable risk stratification. Four readily obtainable pre-treatment variables (i.e., any baseline swallowing impairment per DIGEST ≥1, BOT primary site, T3–4 stage, and age ≥60) reliably identified patients more likely to develop clinically consequential RAD. These predictors align with variables consistently implicated in prior NTCP models for dysphagia and tube feeding dependence^15,31,32^. The value of the present model is that it shifts the prediction target from an arbitrary cross-sectional timepoint to considering the clinical course longitudinally. In resource-constrained settings, such a model could support prioritization or de-prioritization for intensified swallowing surveillance, prophylactic therapy, nutritional intervention, and counseling before treatment begins. The current model was intentionally designed for clinical scalability, prioritizing variables available at the time of treatment planning. Future iterations integrating factors demonstrated to modulate RAD, including radiation dose to swallowing-related structures^14,33–35^, host susceptibility markers such as skeletal muscle mass and physiologic reserve^36,37^, genetic determinants of fibrotic response^38,39^, and acute toxicity indicators such as feeding tube dependence during RT^40–42^, may enable finer Phenotype discrimination and more accurate pre-treatment risk stratification.

More broadly, we propose longitudinal RAD Phenotypes as an alternative meaningful outcome framework to single-timepoint toxicity endpoints for survivorship research. Conventional approaches reduce swallowing outcomes to whether dysphagia is present at a predefined follow-up interval, implicitly assuming that patients with equivalent toxicity grades share similar clinical courses. Trials with swallowing as a primary endpoint could use phenotype membership to capture the full arc of recovery rather than a cross-sectional snapshot. To facilitate adoption of Phenotypes as study outcomes, the model parameters and implementation code have been made publicly available alongside eligibility criteria and recommendations for local validation prior to application in independent datasets, with external validation in independent cohorts planned as a next step.

A few limitations warrant acknowledgement. DIGEST is an ordinal measure and treating it as continuous in the HLME model assumes approximately proportional inter-grade differences, an approximation standard in trajectory modeling of ordered outcomes but not strictly met. Our primary goal was to capture overall severity trends rather than grade-to-grade transitions, and the large sample size supported stable parameter estimation under this specification. The HLME model incorporates only DIGEST as the longitudinal outcome and time as a predictor; future iterations integrating dosimetric parameters, host susceptibility factors, and dynamic treatment indicators may improve trajectory discrimination and phenotype prediction. Neither the longitudinal Phenotype model nor the exploratory baseline prediction model has undergone external validation; prospective replication in independent cohorts (including populations treated with different RT modalities, dose constraints, or swallowing care protocols) represents the essential next step before broader clinical adoption. The 30-month observation window incompletely characterizes late-emerging or progressive dysfunction; future work will extend follow-up to characterize late trajectories.

In conclusion, latent trajectory modeling revealed four clinically and biologically distinct RAD Phenotypes that are predictable from pre-treatment characteristics, supporting a shift toward longitudinal phenotyping as an outcome framework for RAD research. In clinical practice, there are ample opportunities for Phenotype-informed, risk-adaptive prehabilitation and survivorship care.

## Data Availability

The analytic code and fitted model parameters required to apply the derived RAD trajectory Phenotypes to external datasets will be openly available in an NIH-supported scientific data repository (Figshare) upon acceptance. Individual cohort data are available upon reasonable request to the corresponding author.

## REFERENCES

1. Lechner M, Liu J, Masterson L, Fenton TR. HPV-associated oropharyngeal cancer: epidemiology, molecular biology and clinical management. Nat Rev Clin Oncol. 2022;19(5):306–327. doi:10.1038/s41571-022-00603-7

2. Ang KK, Harris J, Wheeler R, et al. Human Papillomavirus and Survival of Patients with Oropharyngeal Cancer. N Engl J Med. 2010;363(1):24–35. doi:10.1056/NEJMoa0912217

3. Cao C, Lee A, Kang JJ, et al. Updated Estimates of Patients With Oropharyngeal Cancer in the US. JAMA Netw Open. 2025;8(10):e2539258. doi:10.1001/jamanetworkopen.2025.39258

4. Lin BM, Wang H, D’Souza G, et al. Long term prognosis and risk factors among HPV-associated oropharyngeal squamous cell carcinoma patients. Cancer. 2013;119(19):3462–3471. doi:10.1002/cncr.28250

5. Shah AH, Nguyen MT, Nguyen SA, Pelic JC, Davidson K, O’Rourke AK. Longitudinal Patterns of Radiation-Associated Dysphagia in Patients With Head and Neck Cancer: A Systematic Review. Head Neck. 2026;48(2):597–623. doi:10.1002/hed.70107

6. Vermaire JA, Raaijmakers CPJ, Monninkhof EM, et al. The course of swallowing problems in the first 2 years after diagnosis of head and neck cancer. Support Care Cancer. 2022;30(11):9527–9538. doi:10.1007/s00520-022-07322-w

7. Bhethanabotla RM, Gulati A, Khalsa IK, et al. Swallowing Function Outcomes in Head and Neck Cancer Survivors Followed by a Long-Term Dysphagia Surveillance Protocol. Head Neck. 2025;47(10):2650–2660. doi:10.1002/hed.28166

8. Galloway TJ, Pugh SL, Ridge JA, et al. Long-Term Feeding Tube Use in Head and Neck Cancer Survivors—An Analysis of Patient- and Treatment-Related Factors. Int J Radiat Oncol. 2025;123(3):703–712. doi:10.1016/j.ijrobp.2025.05.064

9. Grant SR, Hutcheson KA, Ye R, et al. Prospective longitudinal patient-reported outcomes of swallowing following intensity modulated proton therapy for oropharyngeal cancer. Radiother Oncol J Eur Soc Ther Radiol Oncol. 2020;148:133–139. doi:10.1016/j.radonc.2020.04.021

10. Hutcheson KA, Nurgalieva Z, Zhao H, et al. Two-year prevalence of dysphagia and related outcomes in head and neck cancer survivors: An updated SEER-Medicare analysis. Head Neck. 2019;41(2):479–487. doi:10.1002/hed.25412

11. Baudelet M, Van den Steen L, Tomassen P, et al. Very late xerostomia, dysphagia, and neck fibrosis after head and neck radiotherapy. Head Neck. 2019;41(10):3594–3603. doi:10.1002/hed.25880

12. Hutcheson KA, Lewin JS, Barringer DA, et al. Late dysphagia after radiotherapy-based treatment of head and neck cancer. Cancer. 2012;118(23):5793–5799. doi:10.1002/cncr.27631

13. King SN, Dunlap NE, Tennant PA, Pitts T. Pathophysiology of Radiation-Induced Dysphagia in Head and Neck Cancer. Dysphagia. 2016;31(3):339–351. doi:10.1007/s00455-016-9710-1

14. Huynh TTM, Dale E, Falk RS, et al. Radiation-induced long-term dysphagia in survivors of head and neck cancer and association with dose-volume parameters. Radiother Oncol. 2024;190:110044. doi:10.1016/j.radonc.2023.110044

15. Pu Y, Yang J, Shui L, Tang Q, Zhang X, Liu G. Risk prediction models for dysphagia after radiotherapy among patients with head and neck cancer: a systematic review and meta-analysis. Front Oncol. 2025;15. doi:10.3389/fonc.2025.1502404

16. Nagin DS, Odgers CL. Group-Based Trajectory Modeling (Nearly) Two Decades Later. J Quant Criminol. 2010;26(4):445–453. doi:10.1007/s10940-010-9113-7

17. Heinolainen A, Nguyen B, Silén S, Renkonen R, Koskinen M. Survival and data-driven phenotypes in head and neck cancer. Sci Rep. 2025;15(1):5985. doi:10.1038/s41598-025-89053-6

18. Shi Q, Mendoza TR, Gunn GB, Wang XS, Rosenthal DI, Cleeland CS. Using group-based trajectory modeling to examine heterogeneity of symptom burden in patients with head and neck cancer undergoing aggressive non-surgical therapy. Qual Life Res Int J Qual Life Asp Treat Care Rehabil. 2013;22(9):10.1007/s11136-013-0380-0382. doi:10.1007/s11136-013-0380-2

19. Christianen MEMC, Verdonck-de Leeuw IM, Doornaert P, et al. Patterns of long-term swallowing dysfunction after definitive radiotherapy or chemoradiation. Radiother Oncol. 2015;117(1):139–144. doi:10.1016/j.radonc.2015.07.042

20. Moreno A, Sahli AJ, Johnson F, et al. Stiefel MD Anderson OroPharynx cancer (MDA-OPC) cohort: a single-institution, prospective longitudinal outcomes study. BMJ Open. 2025;15(11):e106845. doi:10.1136/bmjopen-2025-106845

21. Hutcheson KA, Barrow MP, Barringer DA, et al. Dynamic Imaging Grade of Swallowing Toxicity (DIGEST): Scale development and validation. Cancer. 2017;123(1):62–70. doi:10.1002/cncr.30283

22. Hutcheson KA, Barbon CEA, Alvarez CP, Warneke CL. Refining measurement of swallowing safety in the Dynamic Imaging Grade of Swallowing Toxicity (DIGEST) criteria: Validation of DIGEST version 2. Cancer. 2022;128(7):1458–1466. doi:10.1002/cncr.34079

23. Rosenbek JC, Robbins JA, Roecker EB, Coyle JL, Wood JL. A penetration-aspiration scale. Dysphagia. 1996;11(2):93–98. doi:10.1007/BF00417897

24. Chen AY, Frankowski R, Bishop-Leone J, et al. The development and validation of a dysphagia-specific quality-of-life questionnaire for patients with head and neck cancer: the M. D. Anderson dysphagia inventory. Arch Otolaryngol Head Neck Surg. 2001;127(7):870–876.

25. List MA, D’Antonio LL, Cella DF, et al. The Performance Status Scale for Head and Neck Cancer Patients and the Functional Assessment of Cancer Therapy-Head and Neck Scale. A study of utility and validity. Cancer. 1996;77(11):2294–2301. doi:10.1002/(SICI)1097-0142(19960601)77:11<2294::AID-CNCR17>3.0.CO;2-S

26. Proust-Lima C, Philipps V, Liquet B. Estimation of Extended Mixed Models Using Latent Classes and Latent Processes: The R Package lcmm. J Stat Softw. 2017;78:1–56. doi:10.18637/jss.v078.i02

27. Nylund KL, Asparouhov T, Muthén BO. Deciding on the Number of Classes in Latent Class Analysis and Growth Mixture Modeling: A Monte Carlo Simulation Study. Struct Equ Model Multidiscip J. 2007;14(4):535–569. doi:10.1080/10705510701575396

28. Manduchi B, Borders J, Alsharawy A, et al. Recalibrating MDADI interpretation for HNC trials using updated clinically important change metrics. Oral Present. Published online 2026.

29. Baudelet M, Van den Steen L, Tomassen P, et al. Very late xerostomia, dysphagia, and neck fibrosis after head and neck radiotherapy. Head Neck. 2019;41(10):3594–3603. doi:10.1002/hed.25880

30. Hutcheson KA, Yuk M, Hubbard R, et al. Delayed lower cranial neuropathy after oropharyngeal intensity-modulated radiotherapy: A cohort analysis and literature review. Head Neck. 2017;39(8):1516–1523. doi:10.1002/hed.24789

31. Willemsen ACH, Kok A, van Kuijk SMJ, et al. Prediction model for tube feeding dependency during chemoradiotherapy for at least four weeks in head and neck cancer patients: A tool for prophylactic gastrostomy decision making. Clin Nutr. 2020;39(8):2600–2608. doi:10.1016/j.clnu.2019.11.033

32. Wopken K, Bijl HP, van der Schaaf A, et al. Development and Validation of a Prediction Model for Tube Feeding Dependence after Curative (Chemo-) Radiation in Head and Neck Cancer. PLoS ONE. 2014;9(4):e94879. doi:10.1371/journal.pone.0094879

33. Duprez F, Madani I, De Potter B, Boterberg T, De Neve W. Systematic Review of Dose–Volume Correlates for Structures Related to Late Swallowing Disturbances After Radiotherapy for Head and Neck Cancer. Dysphagia. 2013;28(3):337–349. doi:10.1007/s00455-013-9452-2

34. Kumar R, Madanikia S, Starmer H, et al. Radiation dose to the floor of mouth muscles predicts swallowing complications following chemoradiation in oropharyngeal squamous cell carcinoma. Oral Oncol. 2014;50(1):65–70. doi:10.1016/j.oraloncology.2013.10.002

35. Roe JWG, Carding PN, Dwivedi RC, et al. Swallowing outcomes following Intensity Modulated Radiation Therapy (IMRT) for head & neck cancer - a systematic review. Oral Oncol. 2010;46(10):727–733. doi:10.1016/j.oraloncology.2010.07.012

36. de Bree R, Meerkerk CDA, Halmos GB, et al. Measurement of Sarcopenia in Head and Neck Cancer Patients and Its Association With Frailty. Front Oncol. 2022;12. doi:10.3389/fonc.2022.884988

37. Kitano M, Yasumatsu R. The impact of sarcopenia in the treatment for patients with head and neck cancer. Auris Nasus Larynx. 2024;51(4):717–723. doi:10.1016/j.anl.2024.05.004

38. Grossberg AJ, Lei X, Xu T, et al. Association of Transforming Growth Factor β Polymorphism C™509T With Radiation-Induced Fibrosis Among Patients With Early-Stage Breast Cancer: A Secondary Analysis of a Randomized Clinical Trial. JAMA Oncol. 2018;4(12):1751–1757. doi:10.1001/jamaoncol.2018.2583

39. Zheng L, Guan Z, Xue M. TGF-β Signaling Pathway-Based Model to Predict the Subtype and Prognosis of Head and Neck Squamous Cell Carcinoma. Front Genet. 2022;13. doi:10.3389/fgene.2022.862860

40. Barbon CEA, Peterson CB, Moreno AC, et al. Adhering to Eat and Exercise Status During Radiotherapy for Oropharyngeal Cancer for Prevention and Mitigation of Radiotherapy-Associated Dysphagia. JAMA Otolaryngol Neck Surg. 2022;148(10):956–964. doi:10.1001/jamaoto.2022.2313

41. Hutcheson KA, Bhayani MK, Beadle BM, et al. Eat and Exercise During Radiotherapy or Chemoradiotherapy for Pharyngeal Cancers: Use It or Lose It. JAMA Otolaryngol Neck Surg. 2013;139(11):1127–1134. doi:10.1001/jamaoto.2013.4715

42. van der Laan HP, Bijl HP, Steenbakkers RJHM, et al. Acute symptoms during the course of head and neck radiotherapy or chemoradiation are strong predictors of late dysphagia. Radiother Oncol J Eur Soc Ther Radiol Oncol. 2015;115(1):56–62. doi:10.1016/j.radonc.2015.01.019

