## Supplementary for "Four longitudinal phenotypes of radiation-associated dysphagia following oropharyngeal radiotherapy: a latent class trajectory analysis"

### SUPPLEMENTARY MATERIAL

**eFigure 1.** Distribution of DIGEST grade across timepoints for overall sample (n=650, MBSs studies: n=2,116)

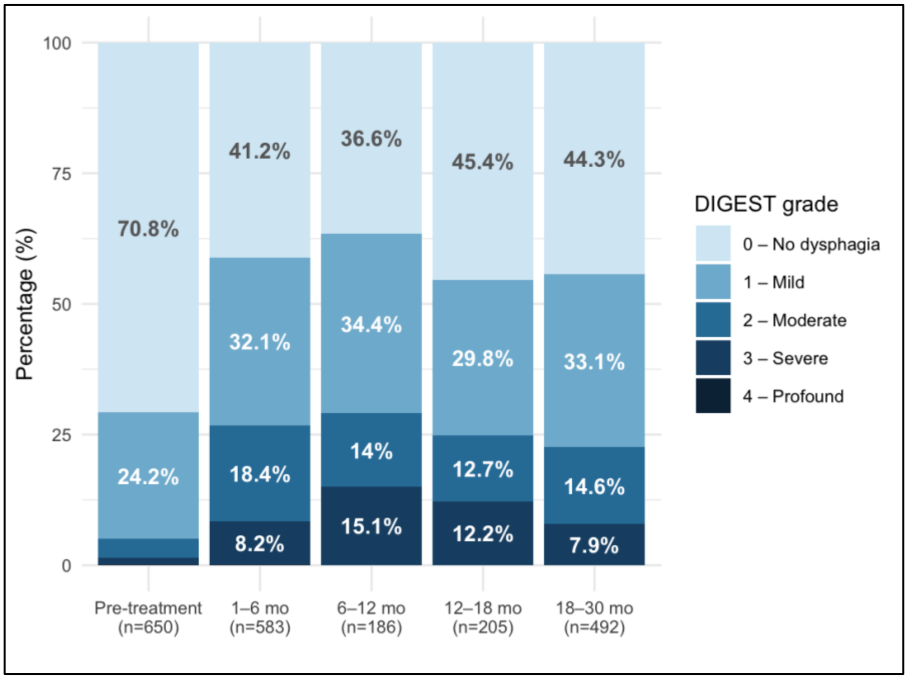

**eTable 1.** HLME model selection across functional forms and number of latent classes

| Functional form | Classes (k) | Log-likelihood | AIC | BIC | Class sizes (%) | Admissible |
|---|---|---|---|---|---|---|
| **Linear** | 1 | −2576.6 | 5161.3 | 5179.2 | 100% | Yes |
| | 2 | −2451.1 | 4918.2 | 4954.0 | 73.1%, 26.9% | Yes |
| | 3 | −2431.7 | 4887.4 | 4941.1 | 70.8%, 13.4%, 15.8% | Yes |
| | 4 | −2417.4 | 4866.7 | 4938.3 | 50.0%, 24.9%, 10.5%, 14.6% | Yes |
| | 5 | −2417.4 | 4874.7 | 4964.2 | — (LogLik identical to k=4) | No |
| **Quadratic** | 1 | −2503.6 | 5017.3 | 5039.7 | 100% | Yes |
| | 2 | −2323.9 | 4667.9 | 4712.6 | 73.1%, 26.9% | Yes |
| | 3 | −2277.3 | 4584.6 | 4651.7 | 70.8%, 13.4%, 15.8% | Yes |
| | 4 | −2243.3 | 4526.6 | 4616.1 | 50.0%, 24.9%, 10.5%, 14.6% | Yes |
| | 5 | −2206.0 | 4462.1 | 4574.0 | 49.4%, 12.2%, 3.7%, 24.8%, 10.0% | No |
| **Natural cubic spline (df=3)** | 2 | −2284.5 | 4593.0 | 4646.7 | 28.8%, 71.2% | Yes |
| | 3 | −2235.5 | 4507.0 | 4587.6 | 68.3%, 10.2%, 21.5% | Yes |
| | 4 | **−2196.3** | **4440.6** | **4548.1** | **59.2%, 16.0%, 14.5%, 10.3%** | **Yes** |
| | 5 | −2144.3 | 4348.6 | 4482.9 | 12.6%, 9.5%, 25.7%, 48.5%, 3.7% | No |
| **Natural cubic spline (df=4)** | 2 | −2279.4 | 4586.9 | 4649.6 | 28.8%, 71.2% | Yes |
| | 3 | −2226.1 | 4494.2 | 4588.2 | 68.9%, 9.4%, 21.7% | No |
| | 4 | −2182.1 | 4420.2 | 4545.6 | 10.6%, 18.6%, 67.1%, 3.7% | No |
| | 5 | −2129.2 | 4328.5 | 4485.2 | 47.2%, 9.1%, 12.8%, 3.7%, 27.2% | No |

*Legend: AIC, Akaike Information Criterion; BIC, Bayesian Information Criterion; df, degrees of freedom; HLME, heterogeneous linear mixed-effects model. Admissibility criterion: all classes must contain ≥10% of the analytic sample. Among admissible models, the model with the lowest BIC was selected (in red)*

**eTable 2**. Fixed effects and classification quality metrics of the longitudinal model for the final 4-class heterogeneous linear mixed-effects model with natural cubic spline trajectory (df=3)

| | Model fixed effects | | | | Sample distribution | | Posterior probability of class assignment | | |
|---|---|---|---|---|---|---|---|---|---|
| **Trajectory Phenotype** | **Term** | **β** | **SE** | **p-value** | **N** | **%** | **Mean** | **Min** | **% above 0.70** |
| **(0) No/Minimal RAD** | Intercept | 0.115 | 0.030 | **<0.001** | 385 | 59.2% | 0.847 | 0.409 | 81.3% |
| | $ns_1$ | 0.044 | 0.105 | 0.675 | | | | | |
| | $ns_2$ | 0.346 | 0.100 | **0.001** | | | | | |
| | $ns_3$ | 0.063 | 0.109 | 0.566 | | | | | |
| **(1) Mild/Moderate RAD** | Intercept | 0.966 | 0.142 | **<0.001** | 104 | 16.0% | 0.776 | 0.373 | 74.0% |
| | $ns_1$ | 0.296 | 0.256 | 0.247 | | | | | |
| | $ns_2$ | 0.291 | 0.308 | 0.345 | | | | | |
| | $ns_3$ | −0.048 | 0.283 | 0.864 | | | | | |
| **(2) Moderate/Severe Transient RAD** | Intercept | 0.348 | 0.073 | **<0.001** | 94 | 14.5% | 0.848 | 0.398 | 77.7% |
| | $ns_1$ | −0.645 | 0.265 | **0.015** | | | | | |
| | $ns_2$ | 3.001 | 0.213 | **<0.001** | | | | | |
| | $ns_3$ | −0.131 | 0.216 | 0.543 | | | | | |
| **(3) Moderate/Severe Progressing RAD** | Intercept | 0.409 | 0.099 | **<0.001** | 67 | 10.3% | 0.890 | 0.523 | 88.1% |
| | $ns_1$ | 2.075 | 0.277 | **<0.001** | | | | | |
| | $ns_2$ | 3.243 | 0.251 | **<0.001** | | | | | |
| | $ns_3$ | 1.192 | 0.218 | **<0.001** | | | | | |

*Note: Posterior probability of assigned class membership ranges from 0 to 1; values ≥0.70 indicate acceptable classification quality.*

*Legend: β, maximum likelihood coefficient estimate; SE, standard error;* $ns_{1–3}$*, natural cubic spline basis functions for months since RT (df=3, automatic knot placement at 3.9 and 13.3 months post-RT).*

**eTable 3**. Cumulative change in predicted DIGEST grade by trajectory class and time period

| **Phase (months post end of RT)** | **(0) No/ Minimal** | **(1) Mild/ Moderate** | **(2) Mod/Severe Transient** | **(3) Mod/Severe Progressing** |
|---|---|---|---|---|
| **Acute** (0–6 mo post-RT) | 0.108 | 0.098 | 1.17 | 0.757 |
| **Subacute** (6–18 mo post-RT) | 0.008 | 0.151 | −0.667 | 1.20 |
| **Late** (18-30 mo post-RT) | 0.044 | −0.222 | 0.480 | −0.172 |

*Note: Values represent the cumulative sum of predicted DIGEST grade change within each time period, derived from model-predicted trajectories*

*Legend: DIGEST, Dynamic Imaging Grade of Swallowing Toxicity; RAD, radiation-associated dysphagia; RT, radiotherapy.*

**eFigure 2.** Observed and model-predicted DIGEST grade trajectories for each latent Phenotype.

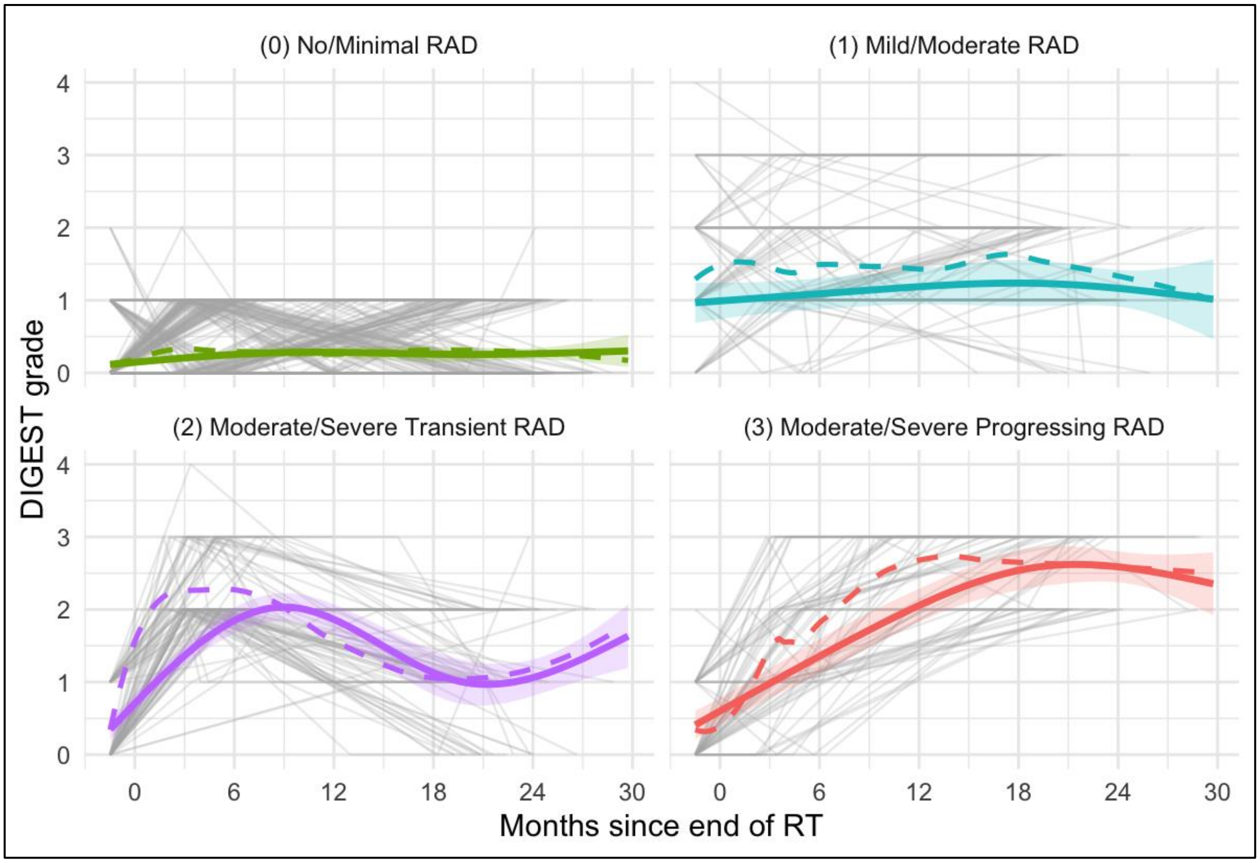


*Note: Individual patient trajectories (grey lines), model-predicted mean (solid line), observed LOESS-smoothed mean (dashed line), and 95% confidence intervals (shaded ribbon) for each of the four latent Phenotype identified by heterogeneous linear mixed-effects modelling with natural cubic spline trajectory (df=3; n=650 patients, 2,116 MBS assessments). Class sizes: (0) No/Minimal RAD, n=385 (59.2%); (1) Mild/Moderate RAD, n=104 (16.0%); (2) Moderate/Severe Transient RAD, n=94 (14.5%); (3) Moderate/Severe Progressing RAD, n=67 (10.3%).*
*Legend: DIGEST, Dynamic Imaging Grade of Swallowing Toxicity; LOESS, locally estimated scatterplot smoothing; MBS, modified barium swallow study; RAD, radiation-associated dysphagia; RT, radiotherapy*

**eTable 3**. Pre-treatment sample characteristics, overall and by Phenotype reported as class distribution within clinical strata (by row %, n=650)

| | Total | (0) No/ Minimal RAD | (1) Mild/ Moderate RAD | (2) Moderate/ Severe Transient RAD | (3) Moderate/ Severe Progressing RAD |
|---|---|---|---|---|---|
| **N (%)** | 650 (100%) | 385 (59.2%) | 104 (16.0%) | 94 (14.5%) | 67 (10.3%) |
| **Age,** mean (SD) | 61.0 (8.5) | 59.2 (8.1) | 63.9 (7.7) | 63.8 (8.6) | 63.3 (9.2) |
| **Sex** | | | | | |
| Female | 72 (11.1%) | 48 (66.7%) | 11 (15.3%) | 8 (11.1%) | 5 (6.9%) |
| Male | 578 (88.9%) | 337 (58.3%) | 93 (16.1%) | 86 (14.9%) | 62 (10.7%) |
| **Tumor site** | | | | | |
| BOT | 322 (49.5%) | 145 (45.0%) | 74 (23.0%) | 56 (17.4%) | 47 (14.6%) |
| Tonsil | 275 (42.3%) | 201 (73.1%) | 25 (9.1%) | 32 (11.6%) | 17 (6.2%) |
| Other | 53 (8.2%) | 39 (73.6%) | 5 (9.4%) | 6 (11.3%) | 3 (5.7%) |
| **HPV/ p16 status** | | | | | |
| Negative | 48 (7.4%) | 23 (47.9%) | 8 (16.7%) | 9 (18.8%) | 8 (16.7%) |
| Positive | 595 (91.5%) | 359 (60.3%) | 95 (16.0%) | 84 (14.1%) | 57 (9.6%) |
| **T stage** | | | | | |
| T0 | 35 (5.4%) | 27 (77.1%) | 4 (11.4%) | 2 (5.7%) | 2 (5.7%) |
| T1 | 190 (29.2%) | 140 (73.7%) | 19 (10.0%) | 16 (8.4%) | 15 (7.9%) |
| T2 | 235 (36.2%) | 144 (61.3%) | 37 (15.7%) | 40 (17.0%) | 14 (6.0%) |
| T3 | 111 (17.1%) | 59 (53.2%) | 16 (14.4%) | 20 (18.0%) | 16 (14.4%) |
| T4 | 79 (12.2%) | 15 (19.0%) | 28 (35.4%) | 16 (20.3%) | 20 (25.3%) |
| **N stage** | | | | | |
| N0 | 52 (8.0%) | 25 (48.1%) | 7 (13.5%) | 15 (28.8%) | 5 (9.6%) |
| N1 | 423 (65.1%) | 285 (67.4%) | 50 (11.8%) | 50 (11.8%) | 38 (9.0%) |
| N2 | 159 (24.5%) | 65 (40.9%) | 44 (27.7%) | 27 (17.0%) | 23 (14.5%) |
| N3 | 16 (2.5%) | 10 (62.5%) | 3 (18.8%) | 2 (12.5%) | 1 (6.2%) |
| **RT type** | | | | | |
| VMAT | 356 (54.8%) | 210 (59.0%) | 61 (17.1%) | 56 (15.7%) | 29 (8.1%) |
| IMRT | 144 (22.2%) | 85 (59.0%) | 21 (14.6%) | 17 (11.8%) | 21 (14.6%) |
| IMPT | 148 (22.8%) | 89 (60.1%) | 21 (14.2%) | 21 (14.2%) | 17 (11.5%) |
| Stereotactic | 1 (0.2%) | 0 (0.0%) | 1 (100.0%) | 0 (0.0%) | 0 (0.0%) |
| 3D Conformal Proton | 1 (0.2%) | 1 (100.0%) | 0 (0.0%) | 0 (0.0%) | 0 (0.0%) |
| **RT dose (Gy)**, mean (SD) [min; max] | 68.2 (3.8) [18.9; 71.1] | 68.2 (3.9) [18.9; 70.7] | 67.6 (4.7) [42.4; 71.1] | 68.6 (2.9) [55.1; 70.4] | 68.8 (2.6) [60.0; 70.8] |
| **RT dose (binary)** | | | | | |
| <66 Gy | 77 (11.8%) | 45 (58.4%) | 16 (20.8%) | 9 (11.7%) | 7 (9.1%) |
| ≥66 Gy | 573 (88.2%) | 340 (59.3%) | 88 (15.4%) | 85 (14.8%) | 60 (10.5%) |
| **Concurrent CT** | | | | | |
| No | 166 (25.5%) | 111 (66.9%) | 25 (15.1%) | 17 (10.2%) | 13 (7.8%) |
| Yes | 484 (74.5%) | 274 (56.6%) | 79 (16.3%) | 77 (15.9%) | 54 (11.2%) |
| **Surgery (primary)** | | | | | |
| No | 593 (91.2%) | 349 (58.9%) | 90 (15.2%) | 90 (15.2%) | 64 (10.8%) |
| Yes | 57 (8.8%) | 36 (63.2%) | 14 (24.6%) | 4 (7.0%) | 3 (5.3%) |
| **MDADI Composite,** mean (SD) [min; max] | 87.8 (13.4) [24.2; 100] | 90.3 (11.5) [24.2; 100] | 80.7 (17.1) [28.4; 100] | 87.5 (12.7) [47.4; 100] | 84.0 (14.8) [44.2; 100] |
| **PSS-HN Diet** | | | | | |
| Non-oral (0) | 2 (0.3%) | 0 (0.0%) | 2 (100.0%) | 0 (0.0%) | 0 (0.0%) |
| Liquid (10-20) | 3 (0.5%) | 0 (0.0%) | 1 (33.3%) | 2 (66.7%) | 0 (0.0%) |
| Pureed/ non/chewable (30-40) | 8 (1.3%) | 2 (25.0%) | 3 (37.5%) | 0 (0.0%) | 3 (37.5%) |
| Restricted solids (50-80) | 65 (10.9%) | 28 (43.1%) | 19 (29.2%) | 11 (16.9%) | 7 (10.8%) |

| | | | | | |
|---|---|---|---|---|---|
| Full, liquid assist (90) | 41 (6.9%) | 16 (39.0%) | 13 (31.7%) | 7 (17.1%) | 5 (12.2%) |
| Full, no restriction (100) | 476 (80.0%) | 313 (65.8%) | 55 (11.6%) | 63 (13.2%) | 45 (9.5%) |
| **Baseline DIGEST** | | | | | |
| 0 | 460 (70.8%) | 344 (74.8%) | 9 (2.0%) | 62 (13.5%) | 45 (9.8%) |
| 1 | 157 (24.2%) | 38 (24.2%) | 66 (42.0%) | 32 (20.4%) | 21 (13.4%) |
| 2 | 24 (3.7%) | 3 (12.5%) | 20 (83.3%) | 0 (0.0%) | 1 (4.2%) |
| 3 | 8 (1.2%) | 0 (0.0%) | 8 (100.0%) | 0 (0.0%) | 0 (0.0%) |
| 4 | 1 (0.2%) | 0 (0.0%) | 1 (100.0%) | 0 (0.0%) | 0 (0.0%) |

*Legend: BOT, base of tongue; CT, chemotherapy; DIGEST, Dynamic Imaging Grade of Swallowing Toxicity; IMPT, intensity-modulated proton therapy; IMRT, intensity-modulated radiation therapy; MDADI, MD Anderson Dysphagia Inventory; PSS-HN, Performance Status Scale for Head and Neck Cancer; RAD, radiation-associated dysphagia; RT, radiation therapy; VMAT, volumetric modulated arc therapy.*
